# Seroconversion rates following COVID-19 vaccination amongst patients with malignant disease- the impact of diagnosis and cancer-directed therapies

**DOI:** 10.1101/2021.05.07.21256824

**Authors:** Astha Thakkar, Jesus Gonzalez Lugo, Niyati Goradia, Radhika Gali, Lauren C. Shapiro, Kith Pradhan, Shafia Rahman, So Yeon Kim, Brian Ko, R. Alejandro Sica, Noah Kornblum, Lizamarie Bachier-Rodriguez, Margaret McCort, Sanjay Goel, Roman Perez-Soler, Stuart Packer, Joseph Sparano, Benjamin Gartrell, Della Makower, Yitz D Goldstein, Lucia Wolgast, Amit Verma, Balazs Halmos

## Abstract

As COVID-19 has been shown to adversely affect patients with cancer, prophylactic strategies are critically needed. We determined the immunogenicity of COVID-19 vaccination in a cohort of cancer patients that had received full dosing with one of the FDA-approved COVID-19 vaccines. 201 oncology patients underwent anti-spike protein SARS-CoV-2 IgG testing post vaccination and demonstrated a high rate of seroconversion (94%) overall. When compared to solid tumors (98%), a significantly lower rate of seroconversion was observed in patients with hematological malignancies (85%), particularly recipients of anti-CD20 therapies (70%) and stem cell transplantation (74%). Patients receiving immune checkpoint inhibitor therapy (97%) or hormonal therapies (100%) demonstrated high seroconversion post-vaccination. Patients with prior COVID-19 infection demonstrated higher anti-spike IgG titers post-vaccination. Relatively lower IgG titers were noted following vaccination with the adenoviral when compared to the mRNA-based vaccines. These data demonstrate generally high immunogenicity of COVID-19 vaccination in oncology patients and identify vulnerable cohorts that need novel vaccination or passive immunization strategies.

## Introduction

COVID-19 can result in increased morbidity and mortality in cancer patients^1,2^ suggesting the need for prophylactic strategies in this vulnerable population. We reported the a large US cohort experience highlighting the significant morbidity and mortality amongst patients with a malignant diagnosis and noted age, co-morbidities and poor performance status as key features leading to a higher risk of adverse outcomes^2^. In addition, we also noted that patients with hematological malignancies and lung cancer carry particularly high morbidity and mortality. These findings subsequently were confirmed in multiple institutional and cohort studies ^3,4^. We then went on to present on seroconversion rates post-COVID-19 infection in a cohort of patients with a malignant disease^5^. Our recent study noted an overall high seroconversion rate as assessed by seropositivity by the SARS-CoV-2 nucleocapsid assay analogous to that of a general patient population with especially high seroconversion rates amongst patients treated with hormonal or immune check-point inhibitor therapies. On the other hand, we observed a significantly lower seroconversion rate amongst patients with hematological malignancies, in particular patients following a stem cell transplantation or anti-CD20 or CAR-T-cell therapies. These results suggested that overall high seroconversion rates might be anticipated following COVID-19 vaccinations as well with likely reduced immunogenicity in certain subgroups of patients suffering from different degrees and mechanisms of immune suppression. Patients with cancer can be immune-compromised due to a multitude of factors, such as the underlying malignancy itself, bone marrow suppressive effects of cytotoxic chemotherapy, prior or ongoing treatments with high degree of immune-suppressive effects, such as corticosteroids, B-cell depleting therapies, i.e. anti-CD20 antibodies, cellular therapies, especially CAR-T cells and stem cell transplantation.

Knowledge as to the immunogenicity of approved vaccines is of critical importance to understand the need of ongoing social isolation and other strategies to mitigate the risk of contracting COVID-19 by vulnerable patients, designing and rapidly conducting clinical studies focused on passive immunization strategies and vaccine trials assessing unique schedules to enable boosting of immune response. However, trials of the currently approved COVID-19 vaccines in general, excluded patients with a diagnosis of a malignancy, therefore information on the safety and efficacy of these vaccines as to the development of effective immunity currently is extremely sparse. Limited published studies and general experience have not identified unique safety signals so far ^6-9^. Given the higher morbidity and mortality of patients with cancer with COVID-19, their ongoing need to be exposed to the health care system and their frequent need for immune suppressive therapies that could further enhance vulnerability to infections, patients with cancer have been identified as a high-priority subgroup for COVID-19 vaccinations-an effort supported by multiple key organizations^10-13^.

While patients with cancer do clearly represent a highly susceptible group with a strong and immediate need to be protected by available, effective vaccines, there remain many uncertainties as to effective vaccinations of patients with cancer. For example, following certain highly immune suppressive therapies-such as an autologous or allogeneic bone marrow transplant, anti-CD20 or T-cell directed regimens vaccinations have low efficacy and their best timing is unclear^14^. Most expert organizations might recommend vaccinations no earlier than 3 months for example following stem cell transplantation or adoptive cell therapies ^15^. Such guidance is lacking for patients needing ongoing immune suppressive therapies and there is a lack of knowledge also as to best approach towards effective vaccination of patients undergoing cytotoxic chemotherapy. One small randomized study did not suggest notable differences as to influenza vaccine immunogenicity efficacy dependent on whether vaccination was given on day of chemotherapy or during the neutropenic period of the treatment cycle ^16^. While many agencies have suggested administering vaccines 1-2 weeks prior to a chemotherapy dose, however this recommendation has not been practical with limited vaccination slot availability, variable chemotherapy (e.g. weekly) and vaccine administration schedules (e.g. 2 doses of Pfizer are recommended to be given 21 while Moderna 28 days apart) leading to liberal recommendations to allow most rapid vaccination of these vulnerable patients^13,17^. Vaccine safety and immunogenicity information is also generally lacking in the context of immune-stimulating therapies, such as checkpoint inhibitor therapy with a few small studies suggesting general safety and of possibly heightened immunity in this context^18^

In order to narrow this key knowledge gap, we set out to comprehensively determine the immunogenicity of FDA approved vaccines in a cohort of patients with a diagnosis of a malignancy via evaluation of rates of anti-spike IgG antibody positivity following vaccination with one of the FDA approved COVID-19 vaccines.

## Methods

### Patient data collection

The study was approved by the Montefiore-Einstein Institutional Review Board. This study was designed as a cross-sectional cohort study and enrolled subjects being seen in the outpatient practices of the Montefiore/Einstein Cancer Center during April 2021. Participants were enrolled in the study after signing informed consent. Subjects underwent SARS-CoV-2 Spike IgG assay, completed a questionnaire focusing on details and side effects of COVID-19 vaccination and provided optional consent for future biobanking for research. The protocol also allowed data collection via retrospective chart review for a small number of patients who underwent SARS-CoV-2 IgG spike antibody testing after vaccination as standard-of-care testing at the discretion of their oncologist.

### SARS-CoV-2 Spike IgG assay

The AdviseDx SARS-CoV-2 IgG II assay was used for the assessment of anti-spike IgG antibody testing. AdviseDx is an automated, two-step chemiluminescent immunoassay performed on the Abbott i2000SR instrument. The assay is designed to detect IgG antibodies directed against the receptor binding domain (RBD) of the S1 subunit of the spike protein of SARS-CoV-2. The RBD is a portion of the S1 subunit of the viral spike protein and has a high affinity for the angiotensin converting enzyme 2 (ACE2) receptor on the cellular membrane^19,20^. The procedure, in brief, is as follows. Patient serum containing IgG antibodies directed against the RBD is bound to microparticles coated with SARS-CoV-2 antigen. The mixture is then washed of unbound IgG and anti-human IgG, acridinium-labeled, secondary antibody is added and incubated. Following another wash, sodium hydroxide is added and the acridinium undergoes an oxidative reaction which releases light energy which is detected by the instrument and expressed as relative light units (RLU). There is a direct relationship between the amount of IgG anti-spike antibody and the RLU detected by the system optics. The RLU values are fit to a logistic curve which was used to calibrate the instrument and expresses results as a concentration in AU/mL (arbitrary units/milliliter). This assay recently has shown outstanding sensitivity (100%) and positive percent agreement with other platforms and demonstrated also high specificity both in the post COVID-19 infection and post vaccination settings. The cutoff value for this assay is 50 AU/mL with <50AU/ml values reported as negative.

### Statistics

Association between two categorical variables was tested with a Fisher exact test. Association between one categorical and one ordinal variable was tested with a Kruskal-Wallis Rank Sum test. Pre-specified hypotheses to be tested included assessing correlation of seropositivity with solid and hematologic malignancies and between the overall cohort and highly immunosuppressive therapies. All analyses were done in R (version 3.6.2).

## Results

### Study cohort

Two hundred and thirteen patients were enrolled in the study via informed-consent process. An additional 29 patients who underwent SARS-CoV-2 Spike IgG testing as standard-of-care were assessed for inclusion in the analysis. Eighteen patients were excluded as they did not have a SARS-CoV-2 Spike IgG test performed at the time of analysis. Another 20 patients were excluded as they had a SARS-CoV-2 spike IgG test before completion of full vaccination series according to FDA guidance (6 with negative and 14 with positive results). Two more patients were excluded who had a negative SARS-CoV-2 Spike IgG and no clear documentation of dates or types of vaccine and one was excluded as data was entered in duplicate. Finally, 201 patients were included in the ultimate safety/immunogenicity analysis who had completed the FDA recommended at least two doses of the mRNA vaccines (Pfizer BNT162b2^21^ or Moderna mRNA-1273^22^) or at least one dose of the adenoviral (Johnson and Johnson (JnJ) AD26.COV2.S^23^) vaccine. Serological data (positive or negative IgG test) from these 201 patients was used in association studies between cancer sub-types and treatments. We also wanted to investigate association between the quantitative titer of SARS-CoV-2 Spike IgG and cancer subtypes and treatments. One hundred eighty-six of 201 patients had IgG titers available who were at least 7 days post the last dose of the vaccine (‘vaccinated cohort’). A population of de-identified patients without a cancer diagnosis who had completed COVID-19 vaccination and received a SARS-CoV-2 IgG spike antibody test > 7 days after their most recent vaccine dose was used as control cohort. (26 subjects at least 7 days post 2-dose mRNA vaccination). This is represented in the CONSORT diagram (**Fig 1**)

**Figure 1:**
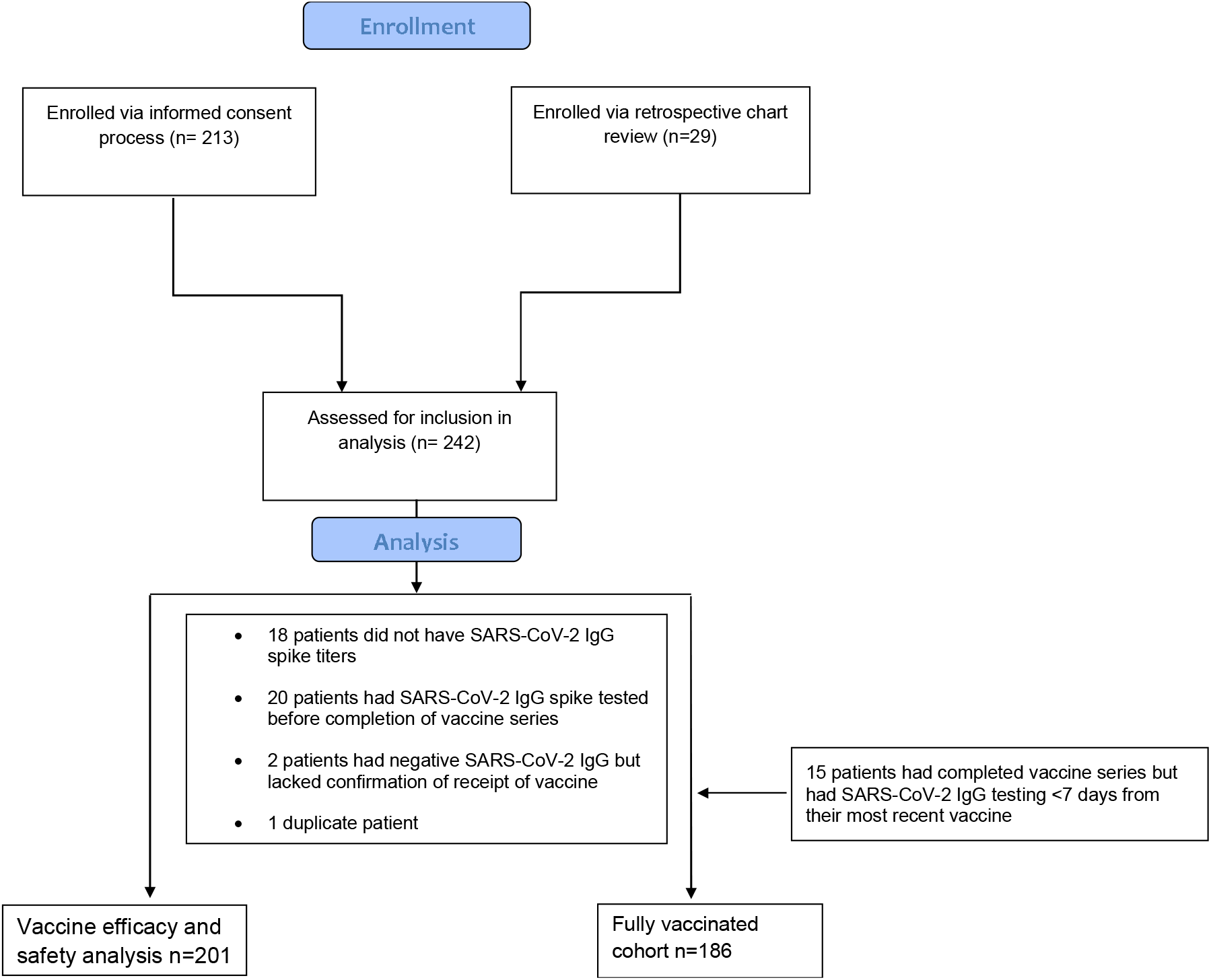
CONSORT diagram showing patient cohort

### Baseline characteristics

A total of 201 patients who completed their vaccinations according to FDA guidance were included in the study. The median age of the patient population was 66 years (range 27-90 years). Fifty-eight percent (117/201) patients were female and 42% (84/201) patients were male. The ethnicity/race of the subjects represented the diverse patient population of the Bronx, NY. 62 (31%) subjects identified their ethnicity as Black, 80 (40%) as Hispanic and 37 (18%) as Caucasian. 134 (67%) of the patients were diagnosed with a solid tumor while 67 (33%) patients had a hematological malignancy with a balanced representation of all common cancer types (**Table 1**). As patients were recruited from our outpatient hematology/oncology clinics, the majority of patients had an active cancer diagnosis. 151 (75%) subjects had an active malignancy and 135 patients (67%) were in active cancer therapy at the time of their vaccination with 113 (56%) of patients on active chemotherapy. 39 (19%) patients were on active chemotherapy within 48 hours of at least one of the vaccine doses. Types of cancer therapies are listed in detail in **Table 2**. 116 patients (54%) had completed vaccination with the Pfizer/BioNTech and 62 (31%) with the Moderna mRNA vaccine and 20 (10%) patients had received the single dose JnJ vaccine. Three patients had received a complete mRNA vaccination series, however the information about the type (Pfizer vs Moderna) was not available.

**Table 1:** Baseline Characteristics.

| **Table 1: Baseline Characteristics** |  |  |
| --- | --- | --- |
| **Age** (Median (Range)) | 67 (27-90) yrs |  |
| **Sex** | **n** | **%** |
| Male | 84 | 42% |
| Female | 117 | 58% |
| **Type of malignancy** |  |  |
| Solid tumor | 134 | 67% |
| Hematologic malignancy | 67 | 33% |
| **Malignancy category** |  |  |
| ***Solid tumor*** | **n** | **%** |
| Breast | 51 | 25% |
| Gastrointestinal | 27 | 13% |
| Genitourinary | 18 | 9% |
| Gynecologic oncology | 10 | 5% |
| Thoracic H&N | 25 | 12% |
| Skin/MSK | 2 | 1% |
| Carcinoma of unknown primary | 1 | 0% |
| ***Hematologic malignancy*** | **n** | **%** |
| Lymphoid | 26 | 13% |
| Myeloid | 18 | 9% |
| Plasma cell | 23 | 11% |
| **Cancer status at the time of vaccine\*** |  |  |
| Active | 110 | 55% |
| Progressive | 7 | 3% |
| Relapse/Recurrent | 34 | 17% |
| Remission | 50 | 25% |
| **Type of vaccine** |  |  |
| Pfizer | 116 | 58% |
| Moderna | 62 | 31% |
| Johnson and Johnson | 20 | 10% |
| mRNA (type unknown) | 3 | 1% |

\*Active cancer: Patient with an initial cancer diagnosis on treatment including surgery, radiation, neoadjuvant, adjuvant or systemic chemotherapy or maintenance therapy (ex- lenalidomide for myeloma or immunotherapy maintenance for non-small cell lung cancer) or not on treatment and under active surveillance
Remission: Patient with cancer diagnosis in the past who has completed cancer-directed therapy and is now only undergoing surveillance.
Relapse/Recurrent: Patient with cancer diagnosis who had completed cancer-directed therapy and achieved remission or was on maintenance therapy now experiencing disease that needs additional treatment
Progressive: Patient with cancer diagnosis who developed disease progression while on first-line systemic therapy

**Table 2.**
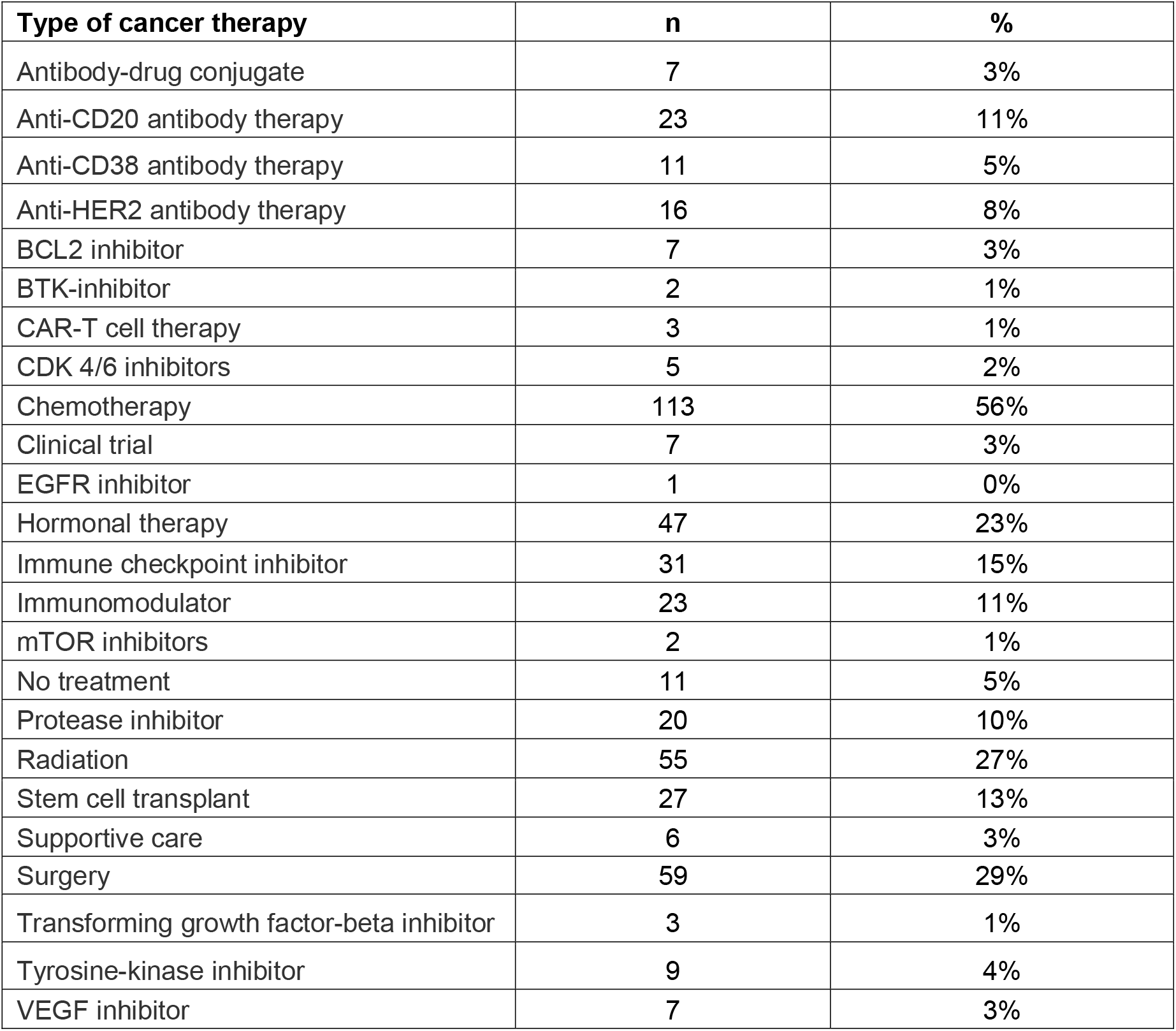
Types of cancer therapy for the cohort.

| Type of cancer therapy | n | % |
| --- | --- | --- |
| Antibody-drug conjugate | 7 | 3% |
| Anti-CD20 antibody therapy | 23 | 11% |
| Anti-CD38 antibody therapy | 11 | 5% |
| Anti-HER2 antibody therapy | 16 | 8% |
| BCL2 inhibitor | 7 | 3% |
| BTK-inhibitor | 2 | 1% |
| CAR-T cell therapy | 3 | 1% |
| CDK 4/6 inhibitors | 5 | 2% |
| Chemotherapy | 113 | 56% |
| Clinical trial | 7 | 3% |
| EGFR inhibitor | 1 | 0% |
| Hormonal therapy | 47 | 23% |
| Immune checkpoint inhibitor | 31 | 15% |
| Immunomodulator | 23 | 11% |
| mTOR inhibitors | 2 | 1% |
| No treatment | 11 | 5% |
| Protease inhibitor | 20 | 10% |
| Radiation | 55 | 27% |
| Stem cell transplant | 27 | 13% |
| Supportive care | 6 | 3% |
| Surgery | 59 | 29% |
| Transforming growth factor-beta inhibitor | 3 | 1% |
| Tyrosine-kinase inhibitor | 9 | 4% |
| VEGF inhibitor | 7 | 3% |

### Overall Immunogenicity and Safety of SARS-CoV2 Vaccine

SARS-CoV2-anti-spike protein antibody test (Abbott) was performed and demonstrated a high rate of seropositivity (94%) with only 13 (6%) subjects with a negative value (titer below 50 AU/ml). Percent positivity appeared similar between the vaccine types (Pfizer 95%, Moderna 94%, JnJ 85% positivity) with a trend noted towards lesser positivity with the JnJ vaccine. We also assessed antibody titers in a sub-cohort of 186 patients with available IgG levels >7 days post vaccination (vaccinated cohort matching the definition of our non-cancer control cohort). Highest titers were seen with the Moderna vaccine (Median 11963 AU/ml, SD 18742) followed by the Pfizer/BioNT vaccine (Median 5183 AU/ml, SD 16621) and the single dose JnJ vaccine (Median 1121 AU/ml, SD 17571) (P Val<0.05, Kruskall Wallis Test, **Fig 2A**).

**Fig 2:**
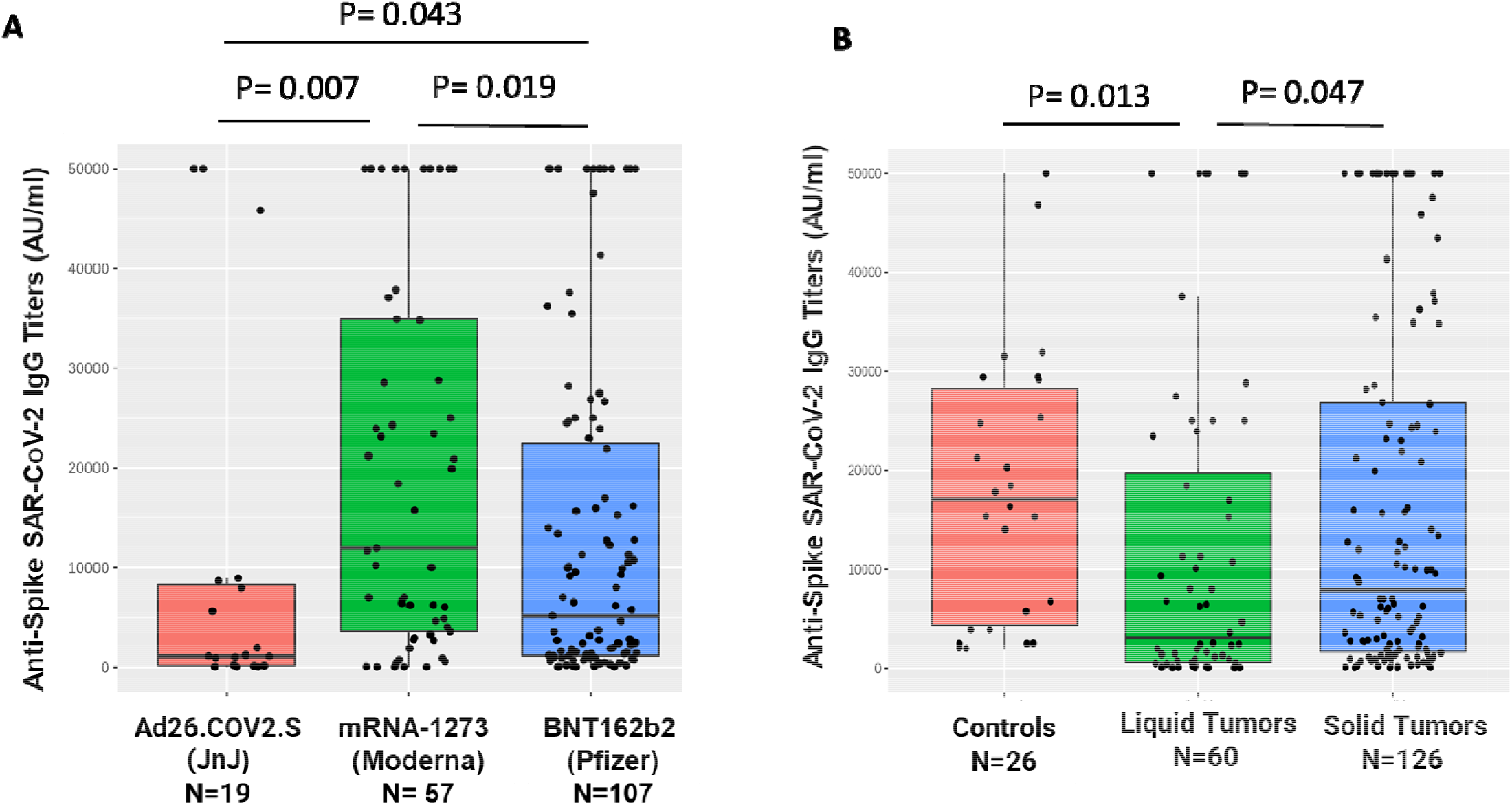
Association of anti-SARS-CoV2 Spike IgG titers with Vaccine Types and cancer Subtypes: Spike protein antibody titers were significantly higher with mRNA vaccines (mRNA-1273 and BNT162b2) when compared to the AD26.COV2.S adenoviral vaccine (A). mRNA-1273 vaccine was associated with the highest titers (A). Patients with liquid tumors h; significantly lower titers when compared to solid tumors and non cancer patient controls. No difference was seen between solid tumor patients and controls. (B). Differences assessed by kruskal Wallis Test.

Overall vaccinations appeared to be generally very safe amongst this cohort with mostly mild and moderate anticipated side effects reported. Sore arm/muscle aches was the most commonly reported adverse event (40% of patients reported it with first dose of vaccine while 34% reported with second dose). No patient required an emergency room visit or hospital admission for side effect management. Sixty-two percent patients reported no side effects with the first and 48% reported no side effects with second dose whatsoever. Only 2% rated their adverse events as severe with each dose.

### Solid tumors versus hematological malignancies

In the cohort of patients with solid tumors, seropositivity post vaccination was found to be extremely high (98%). while a significantly lower seropositivity rate was seen in the cohort of patients with a hematological malignancy (85%, p=0.001 by Fisher’s exact test). Analysis of a sub-cohort of 186 patients with available IgG titers >7 days post vaccination revealed analogous statistically significant differences with significantly higher titer values in solid tumors (median 7858AU/ml, SD 18103) when compared to liquid tumors (median 3068AU/ml, SD 16540, p=0.013 by Kruskal-Wallis). Comparison of titers from a cohort of controls (26 patients who had completed 2 doses of mRNA based vaccinations >7days prior to testing) revealed no significant difference when compared to solid tumor patients but showed a statistically significant difference when compared to the cohort of patients with hematological malignancy (p=0.01, **Fig 2B**). Seropositivity rates amongst control were 100%.

### Association with active cancer therapies and highly immune suppressive therapies

No significant differences in seroconversion were seen in when comparing patients on active cancer therapy versus patients who were not (95% vs 93%). However, significantly lower rates of seropositivity were seen in the cohort of patients on active cytotoxic chemotherapy (89%) versus others (99%, p=0.007) without notable differences in titer levels (**Suppl Fig 1**). Next, we focused our analysis on key cohorts of patients who had received highly immune suppressive therapies, such as stem cell transplant, anti-CD20 therapy or CAR-T cell therapy. We observed significantly lower seroconversion rates in all these pre-defined special interest subsets – prior stem cell transplant (74%, p=0.004), anti-CD20 therapies (70%, p=0.0001) and CAR-T cell treatments (all 3 patients remained seronegative after vaccination, p=0.0002) (**Table 3**). Of the 27 stem cell transplant patients, 24 received an autologous and 3 an allogeneic transplant (2 seropositive, one seronegative). Accordingly, significantly lower titer levels were also seen in patients receiving anti-CD20 therapies compared to the overall group of patients (**Fig 3**). These results highlight the continued vulnerability of such patients during the pandemic.

**Table 3:**
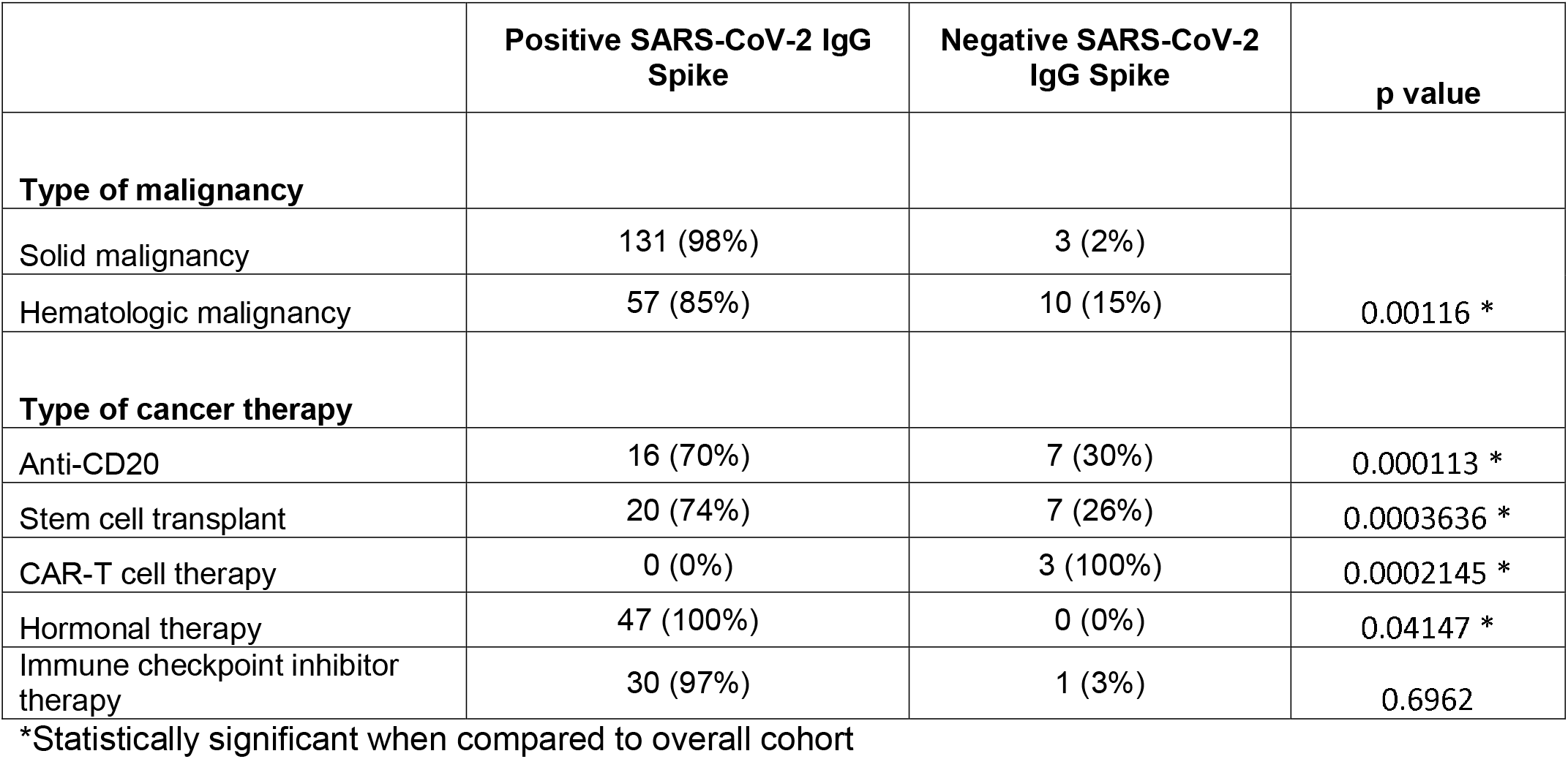
Association of Anti-Spike IgG with Patient and disease characteristics.

**Fig 3:**
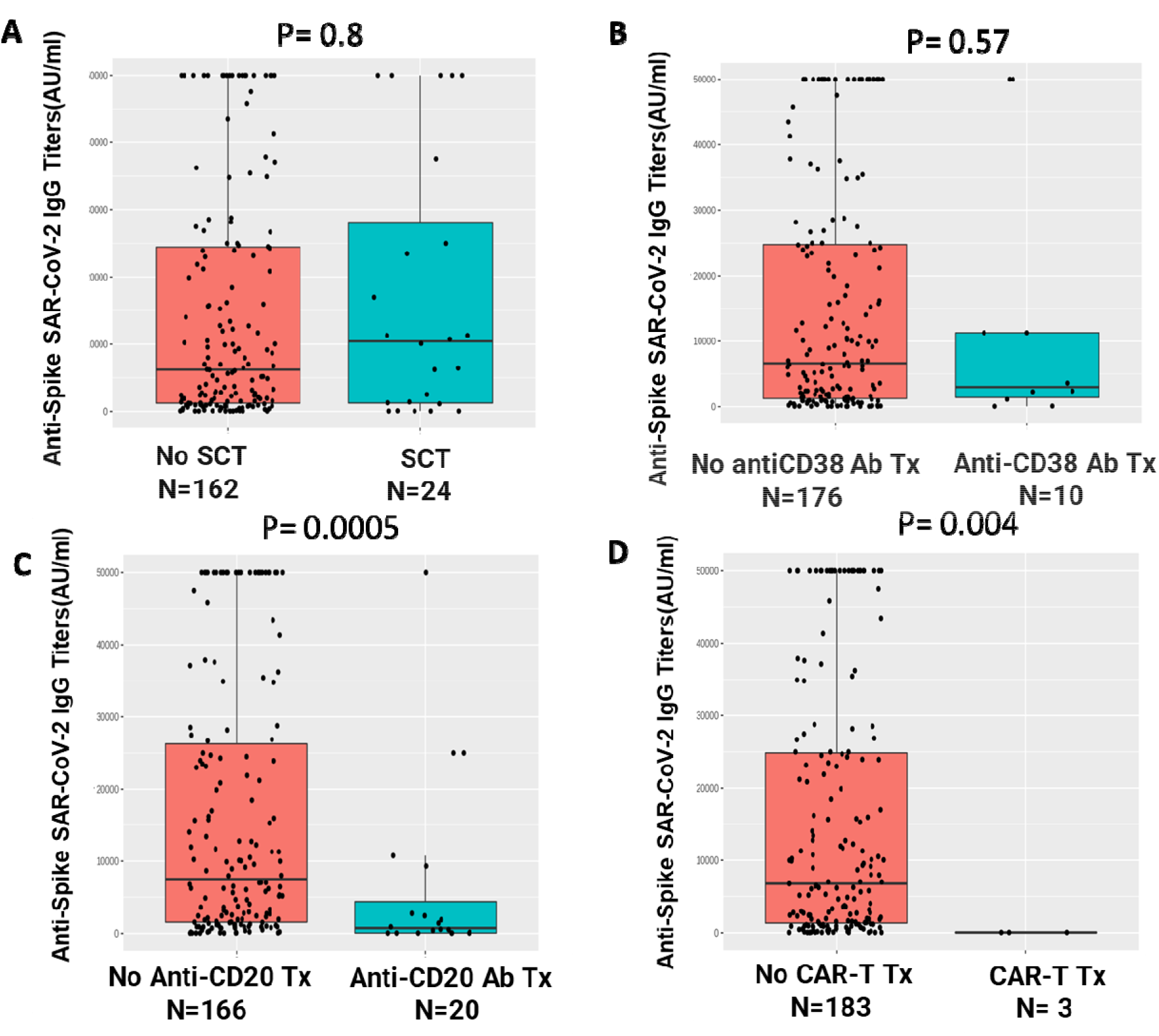
Association of anti-SARS-CoV2 Spike IgG with lmmuno-suppressive therapies: Spike protein antibodies after full vaccination did not significantly differ in patients receiving stem cell transplantation (SCT)(A) and Anti-CD38 antibody therapy (B) when compared to respective counterparts. Patients receiving Anti-CD20 antibody treatments and Chimeric antigen receptor T (CAR-T) therapy had a significantly lower titer after vaccination (C,D). Differences assessed by Kruskal Wallis Test.

### Association with additional oncological therapies

We observed high rates of seroconversion post vaccination in patients on hormonal therapy (100% seropositivity, p=0.04) and checkpoint inhibitor therapy (97%). Interestingly, while all patients on CDK4/6 inhibitor treatment showed positive anti-spike IgG test results, notably antibody titers were very low in this small subset (median 930AU/ml SD 496 versus median 6637AU/ml, SD 17710 for overall cohort) (**Fig 4**). Given known involvement of the CDK4/6 pathway in immune activation^24^ this might be biologically plausible and calling for further studies into this subset. We also noted trends towards lower titers amongst other subgroups, such as patients having received BCL2 or BTK-directed therapy in line with prior observations as to vaccine efficacy ^25^ (**Suppl figure 2**).

**Fig 4:**
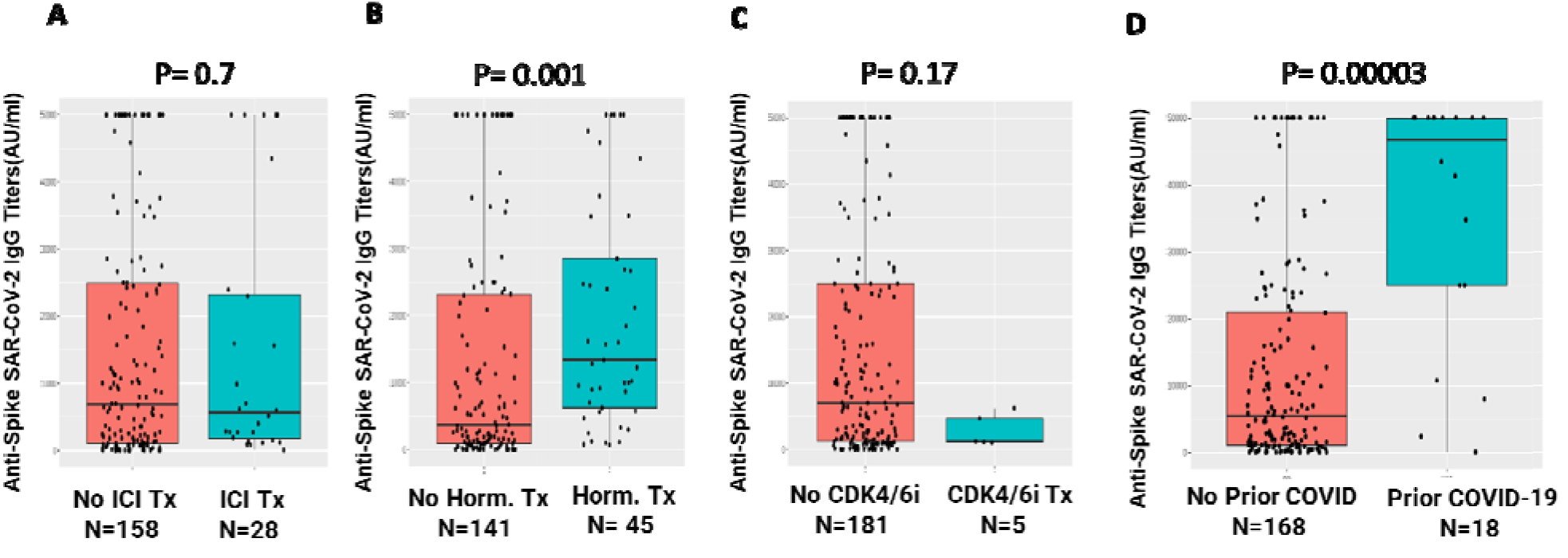
Association of anti-SARS-COV2 Spike IgG titers with type of therapy and prior COVID-19 infection: Spike protein antibody titers after full vaccination did not significantly differ in patients receiving Immune checkpoint Inhibitor Theraphy (ICI) (A) when compared to respective counterparts. Patients receiving hormonal therapy had a significantly higher titer after vaccination (B). Patients receiving CDK4/6 inhibitor therapy had a trend towards lower titer after vaccination (C) Patients with prior COVID-19 infection had significantly higher IgG titers (D). Differences assessed by Kruskal Wallis Test.

### Association with Prior Covid infection

Previous studies have noted heightened antibody responses to COVID-19 vaccinations in patients with a prior COVID-19 infection^26^. Our cohort included 22 subjects with a known prior COVID-19 infection and a high rate of seroconversion was seen in this subset (21/22 seroconverted for a 95% seroconversion rate with sole subject not seroconverting having received an auto-stem cell transplant). Antibody titers in the subgroup with available titers >7 days post vaccination for the previously infected cohort similarly were strikingly elevated and significantly higher than the overall group (prior COVID-19-median 46737AU/ml, SD 18681; others median 5463AU/ml, SD 16144, p<0.001 by Kruskall Wallis test, **Fig 4D**).

## Discussion

COVID-19 disease has had a devastating impact worldwide and especially so among patients with a cancer diagnosis. Patients with cancer are known to be particularly predisposed to adverse outcomes with SARS-Cov-2 infection related to a number of issues such as impact of underlying disease on performance status, age/co-morbidities of affected patients, immune suppression related to disease such as in patients with hematological malignancies as well as immune suppressive effects of disease-directed therapies^27-30^. In addition, patients with cancer requiring active therapy face frequent exposure to the health care system increasing risk of acquiring COVID-19. Lastly, treatment modifications due to the ongoing pandemic can compromise disease outcomes amplifying the urgent need to implement widespread vaccination of patients with malignant disease-an initiative with broad support from a large swath of cancer care/advocacy organizations^13^.

While these vaccination efforts must represent highest priority, understanding the immunogenicity of the currently available vaccines amongst patients with cancer dependent on underlying disease/treatment correlations is also of paramount importance as certain subsets of patients- for example patients with a high level of immune suppression are known to respond poorly to vaccinations and knowledge is also lacking as to optimal timing of vaccinations in relationship to ongoing therapy. A cohort study from our institution indeed demonstrated a very high level of seroconversion as measured by nucleocapsid antibody testing in patients with cancer following COVID-19 infection. However, seroconversion rates logically were less amongst patients with a hematological malignancy with notable low seroconversion rates following prior highly immune suppressive therapy, such as stem cell transplantation, anti-CD20 therapy or CAR-T cell therapy while very high seroconversion rates were found amongst patients on hormonal or immunotherapies, such as checkpoint inhibitor therapy^5^.

While the FDA approved vaccines, such as the mRNA based Moderna and Pfizer/BioNTech vaccines and the adenovirus-based Johnson & Johnson vaccines yield high level of protection against the presently circulating variants in the general population, very limited data is currently available as to the immunogenicity of these vaccines as measured by anti-spike IgG antibody positivity following vaccinations by the FDA-approved vaccines amongst patients with a cancer diagnosis. Small cohort studies so far suggest alarmingly low seroconversion rates following a single dose of mRNA vaccination from the UK and France^6,31^ and low seropositivity rates in certain highly susceptible cohorts of patients, such as patients with CLL and myeloma^9,32,33^.

Hereby we present a large cross-sectional cohort study from our institution serving an ethnically diverse patient population. This cohort of 201 patients with fair representation of malignancy subtypes and prior therapies and with completed COVID-19 vaccinations according to FDA guidelines demonstrates encouragingly, a very high 93% seropositivity rate. In line with our prior observations post COVID-19 infection, seroconversion rates were notably lower amongst patients with a hematological malignancy (85% versus 98% in patients with a solid tumor diagnosis). Furthermore, significantly lower seroconversion rates were found in patients following prior stem cell transplant (74%), anti-CD20 therapies (70%) and CAR-T cell therapies (all tested patients remained negative). These findings are biologically meaningful in light of severe long lasting immunologic defects following stem cell transplantation and B-cell depleting therapies especially anti-CD20 antibody therapy, and even more pronounced immune suppression, including hypogammaglobulinemia requiring intravenous immune globulin replacement, following CAR-T cell therapy directed against CD19. Conversely, patients on hormonal (100%) and immunotherapies (97%), such as checkpoint inhibitor therapy had exceedingly high seropositivity rates. Intriguing findings requiring further confirmation in larger cohorts suggest lower antibody titers in patients on CDK4/6 inhibitor therapy-a biologically plausible observation given the role of CDK4/6 in immune cell proliferation^24^. Low titers were also seen amongst patients on BTK inhibitor and Bcl-2 inhibitor directed therapies^9^. In addition, further examination of our cohort confirmed significantly higher titer levels reached following prior COVID-19 infections and also suggests differences as to the immunogenicity of the different types of vaccines with significantly lower titers reached after vaccination with the JnJ vaccines versus the mRNA vaccines.

Several shortcomings of our study need to be listed. These do include limited representation of some patient cohorts not allowing clear conclusions as to seroconversion rates amongst less common malignancy types or less frequently used treatment approaches. However, we believe that based on our data falling fully in line with scientific rationale, such projections for those subsets can logically be made while more focused studies are being conducted. Our cohort also over-represented patients on active therapy as recruitment occurred over a short period in our outpatient departments. In addition, our study analogous to other studies published so far relies solely on seroconversion as defined by anti-spike protein IgG levels as a surrogate for immunity to COVID-19. Admittedly, the anti-spike IgG antibody used in our study, albeit specific to the receptor binding domain (RBD) of the spike protein, might still not necessarily correlate with “neutralizing” activity. In addition, the contribution of T cells, such as effector memory T cells to long-term immunity against COVID-19 is still being defined ^34,35^. Despite these constraints, on a population basis the higher presence of positive anti-spike antibodies in a group of patients is expected to correlate with more neutralizing antibody activity versus another group that has a lower percentage of anti-spike positivity, providing credibility to our data.

Another potential limitation is under-estimation of titer values for anti-spike antibodies as evidence suggests titers may rise over time^36^, however, a cut-off of 7 days was used to match the control cohort and eliminate bias in the analysis.

Our study along with other emerging data strongly highlights the continued need to invest strong efforts to vaccinate patients with a cancer diagnosis urgently and broadly as vaccinations are likely to be highly effective. On the other hand, our study highlights vulnerable cohorts of patients, in particular patients with hematological malignancies following receipt of highly immune suppressive therapies- stem cell transplant, anti-CD20 therapies, CAR-T cell treatments^37^These cohorts of patients will need unique considerations and urgent plans to novel vaccination strategies as well as possibly passive protection strategies in the face of the ongoing pandemic- such as protective antibody therapies.

In summary, we present a large cohort of patients with malignancy who underwent full COVID-19 vaccination according to FDA established guidance. In this cohort of ethnically diverse patients with broad representation of a wide range of malignancies and therapies, encouragingly very high seropositivity rates were observed in contrast from previously published small cohort studies focused on unique subsets of susceptible patients or non-standard vaccination schedules. Statistically significantly lower seropositivity rates were observed in patient cohorts with a high level of immune suppression and these cohorts will warrant innovative strategies for optimal protection. Our findings in general are in support of broad and urgent COVID-19 vaccinations in line with FDA guidance for proper protection of patients with a cancer diagnosis and allowance of optimal cancer treatment delivery in the face of the ongoing COVID-19 pandemic.

## Data Availability

Data is available with the corresponding author upon request

---

**Supplementary figure 1.**
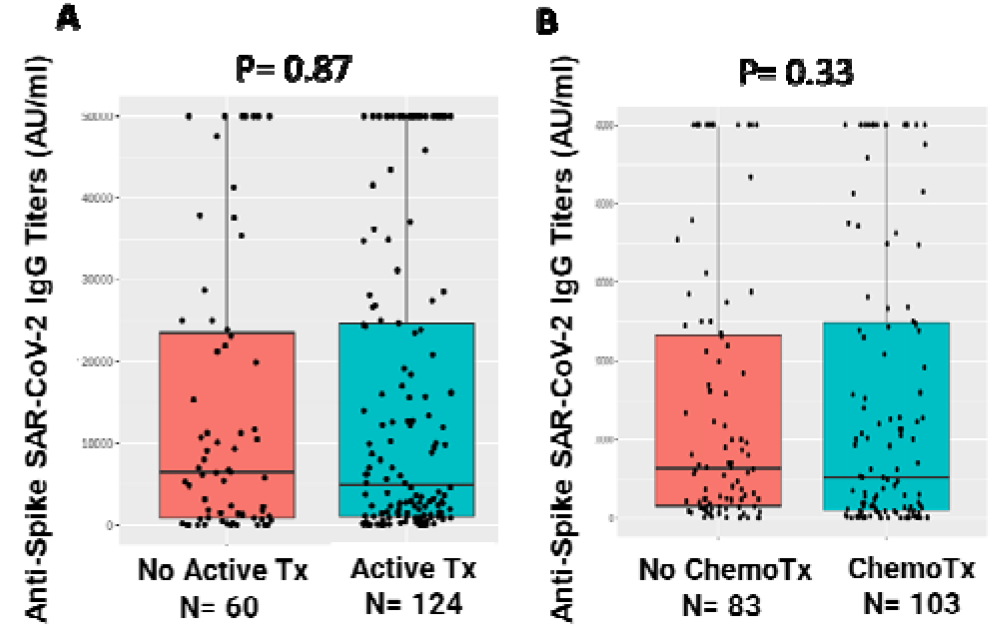
Association of anti-SARS-CoV2 Spilce IgG with active systemic therapy and chemotherapy: Spike protein antibody titers after full vaccination did not significantly differ in patients receiving active therapy (A) or active chemotherapy (B) when compared to respective counterparts. Differences assessed by Kruskal Wallis Test.

**Supplementary figure 2.**
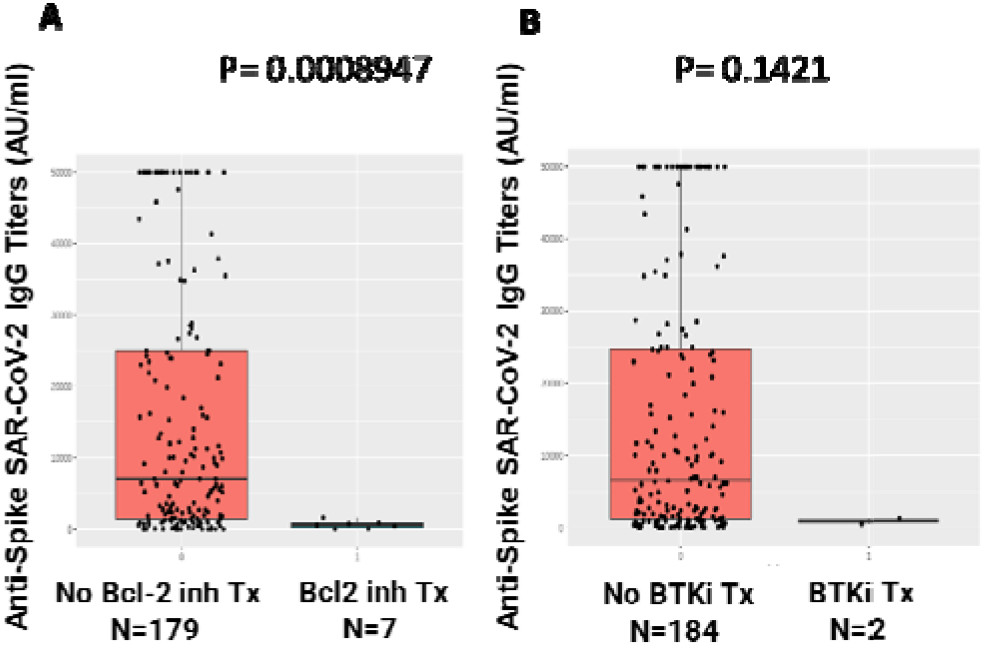
Association of anti-SARS-CoV2 Spilce IgG with systemic therapy: Spike protein antibody titers after full vaccination showed significantly lower titers with Bcl-2 inhibitor venetoclax therapy (A) and trend towards reduced titers with BTK inhibitor therapy as compared to respective counterparts. Differences assessed by Kruskal Wallis Test.

